# Toward AI-Guided Smoking Cessation: Individualized Nicotine Addiction Modeling Using Gaussian Processes

**DOI:** 10.1101/2023.10.25.23297563

**Authors:** Anirudh Chari

## Abstract

Cigarette smoking remains the leading cause of preventable disease and death in the United States, accounting for nearly half a million deaths annually. Given the recent rise of artificial intelligence in healthcare applications, computational assessment of smoking behaviors is a promising direction. In this study, we aim to recognize and classify addiction patterns in individual smokers’ daily usage based on time series data. To this end, we leverage Gaussian process modeling to iteratively learn a function that defines a smoker’s behavior as usage data is accumulated. Namely, we aim to learn weekly periodic trends in usage, and then utilize the model to predict future trends. We demonstrate that the outputted predictions resemble the actual data well, and that these informed forecasts significantly outperform those of a naive prediction model with respect to accuracy. Finally, we propose strategies for utilizing these predictions for goal-setting as part of a computer-supervised gradual cessation program.

## 1 Introduction

Smoking cessation generally comes in two forms: abrupt and gradual. Abrupt cessation has shown to be more effective in maintaining long-term abstinence than self-supervised gradual cessation [1]. However, computer-supervised gradual cessation (i.e., goal-setting algorithms) has not been widely explored, and a preliminary approach showed significantly higher long-term abstinence rates than both abrupt and self-supervised gradual cessation attempts, at least in adolescents [2]. With the recent rise and great success of artificial intelligence (AI) in healthcare applications [3], this is a promising direction.

Modeling drug use and addiction computationally has been widely explored in recent years [4]. Popular approaches include reinforcement learning [5] and dopamine-based modeling [6]. In [7], the author demonstrates that Bayesian frameworks can be effective for analyzing decision-making behaviors in drug addicts. In [8], the authors develop a computational model of nicotine addiction that classifies the severity of a user’s addiction based on neurophysiological indicators.

Time series forecasting is the problem of predicting future data points or trends in a time series, given a segment of the current data. Popular time series forecasting techniques include exponential smoothing, ARIMA models, Kalman filters, long short-term memory (LSTM) models [9], and Gaussian processes (GPs) [10]. Because they operate on Bayesian inference, GPs pose the additional advantage of uncertainty quantification in outputted predictions, which can be especially helpful in evaluating their efficacy in the proposed setting. For an intuitive tutorial on the mechanics of GPs for solving regression problems, see [11].

The main challenge in forecasting is to learn the underlying patterns within potentially noisy data. This makes time series forecasting a very valuable tool in healthcare, as exemplified by its application to disease diagnosis and prognosis [12, 13]. Time series analysis and forecasting have also been applied in modeling large-scale nicotine use and cessation [14], especially in evaluating interventions [15, 16]. However, applying forecasting methods in behavior assessment of individual smokers has not been widely studied in recent years.

We propose that the nicotine use of smokers with respect to time can be modeled using GPs, and that this model can be employed for effective classification and prediction of addiction and cessation behaviors. Ultimately, we aim to leverage these insights to develop a personalized and adaptive gradual smoking cessation program.

## 2 Methods

### 2.1 Preliminary: Gaussian Processes

#### 2.1.1 Definition

A Gaussian process (GP) model describes a probability distribution over all possible functions that fit a set of points. A GP model leverages Bayesian inference to update the posterior function function, defined as the outputted means with the variances as quantified uncertainties, as new data is obtained. This posterior function can be used as a regression model to make predictions about new data.

#### 2.1.2 Kernels

The *kernel* of a GP model defines the general curve fitting behavior, i.e. our foundational beliefs on how our function should behave. More formally, a kernel *k*_*θ*_(*x*_1_, *x*_2_) defines the covariance between two function values *x*_1_ and *x*_2_ using some hyperparameters *θ*. Multiple kernels can be combined through *kernel composition* in order to fit more complex functions.

#### 2.1.3 Hyperparameters

Being the coefficients of our kernel function, the hyperparameters *θ* determine the exact curve fitting behavior of the GP model. Estimating hyperparameters manually is difficult; a common approach is Bayesian optimization.

### 2.2 Modeling Smoking Habits

We observe that weekly patterns emerge in smokers’ nicotine use [17]. Thus, we can formulate the problem as periodical time forecasting, for which GPs have proven to be effective [18]. At a high level, the role of the GP is to learn a function that defines the user’s behavior. Our approach uses the framework provided in [18] as a foundation, and we summarize the framework as it relates to our work below.

#### 2.2.1 Kernel Composition

We utilize a kernel *K*, which is composed by

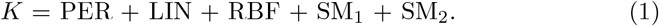

Here, the periodic kernel PER provides a weekly seasonality, the LIN kernel provides a linear trend, and the RBF, SM_1_ and SM_2_ kernels are collectively used to represent nonlinear trends. See [18] for explicit definitions of each of these kernels.

#### 2.2.2 Hyperparameter Estimation

While the model aims to learn the specific parameters defining an individual’s behavior, it also consists of hyperparameters *θ* defining general behavior that applies to the entire function space, i.e., all users’ behaviors. We begin by assigning log-normal priors to each hyperparameter, and then improve our estimations by training the model on various sample data using an iterative process called *maximum a posteriori* (MAP) estimation, as outlined in [18].

#### 2.2.3 The Forecasting Problem

GP regression modeling is naturally applicable to time series forecasting problems. We can formally represent our addiction forecasting task as a regression problem

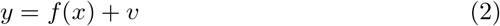

where *y* is nicotine dose per day (measured in number of cigarettes), *x* is time in days, *f* ∼ *GP* (0, *k*_***θ***_) is the function with the GP as a prior distribution, and 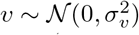 is the noise with variance 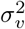. Then, given *m* points of training data *x* = (*x*_1_, …, *x*_*m*_), *y* = (*y*_1_, …, *y*_*m*_), and given *n* test inputs 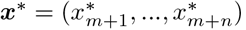, the forecasting problem is to compute the function’s posterior distribution 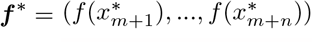.

## 3 Experiments

### 3.1 Dataset

We utilize the dataset provided in [17], which contains a sample of 62 participants who have been smoking for more than two consecutive years and who smoke more than five cigarettes a day. Participants were male and female college students between 18 and 26 years of age. Each participant recorded the number of cigarettes he or she smoked each day for up to 12 consecutive weeks. Some data was incomplete, so we consider only the participants who recorded data for the full 12-week period, which yields *T* = 50 time series. For each time series, we form our training set using the first *m* = 56 days and our test set using the last *n* = 28 days. We normalize each time series to have a mean of 0 and a standard deviation of 1.

### 3.2 Baseline

We compare the performance of the GP model to that of a baseline naive model. The naive model uses the average value of the historical data as its predicted value for all future days. More formally,

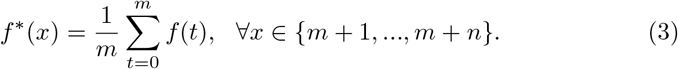

The naive model makes an educated guess that smoking behaviors remain constant, but it leverages no knowledge of weekly patterns in behavior.

### 3.3 Evaluation Metric

The *mean absolute error* (MAE) of the test set is given by

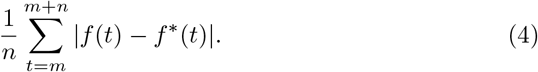

Lower values of MAE indicate better model performance. MAE is a common metric for evaluating regression models on normalized data.

### 3.4 Results

The MAE values across the time series were significantly lower for the GP model (*M* = 0.823, *SD* = 0.168) than for the naive model (*M* = 0.908, *SD* = 0.150), *t*(49) = 4.78, *p <* .001. The GP model successfully discerns a wide variety of periodicities (Fig. 1), and the predicted values and associated error bounds resemble the actual data well, *χ*^2^(26, *N* = 50) = 767.65, *p <* .001.

**Figure 1.**
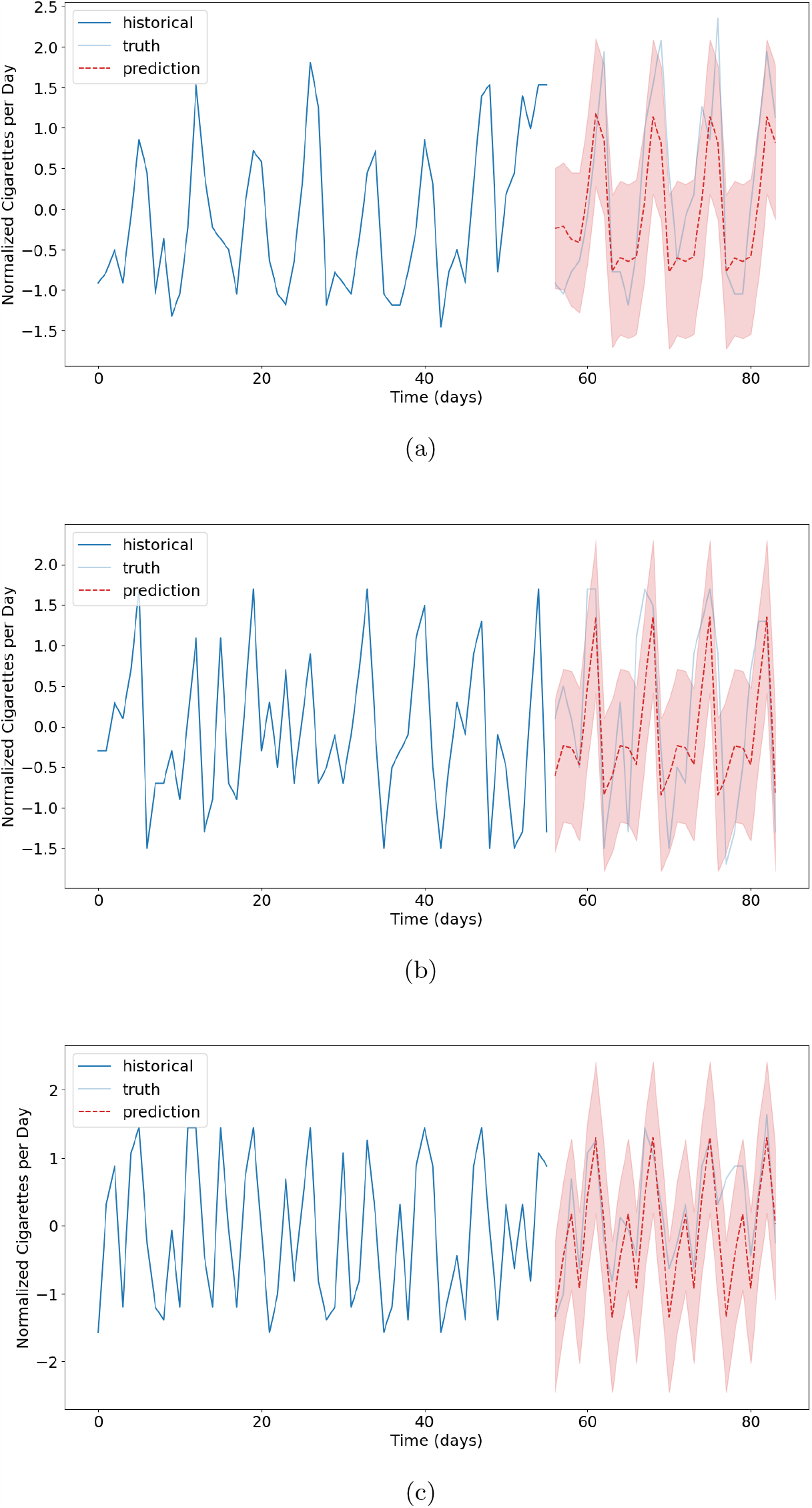
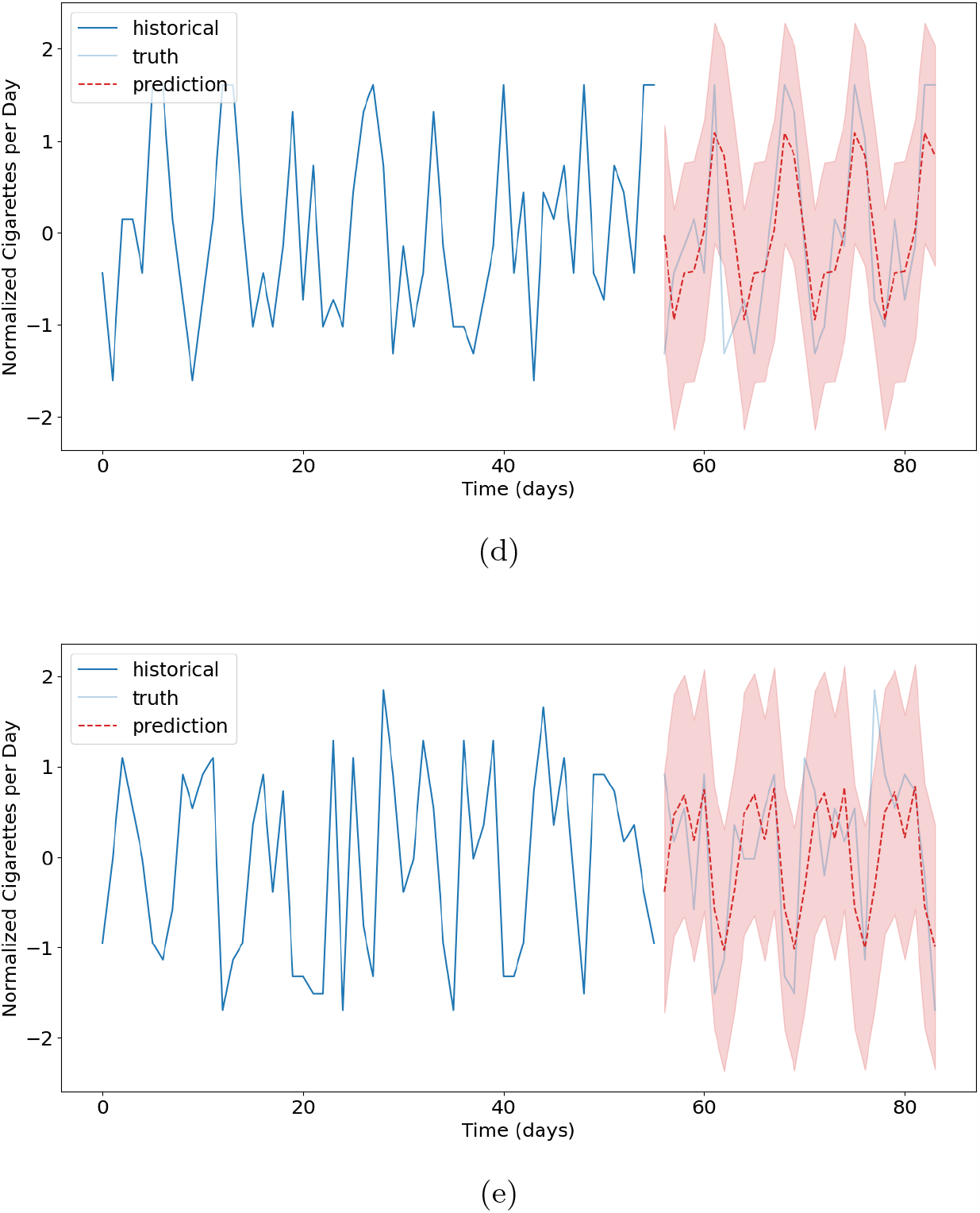
Example outputs from GP forecasting.

## 4 Guided Cessation

We have shown that GPs are an effective tool for modeling nicotine addiction behaviors. We expect that the model will be useful in designing a computer- supervised gradual smoking cessation program. However, this requires an additional goal-setting procedure that actually leverages the model’s insights. Here, we outline some suggested procedures.

### 4.1 Naive Bounding Procedure

At time *t*, let 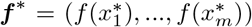 denote the posterior distribution of *f* ∼ *GP* (0, *k*_***θ***_), i.e. the regression output up to *x* = *x*_*m*_ where *t < x*_*m*_. Then a simple goal-setting function is

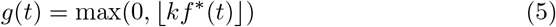

for some constant *k* ∈ (0, 1) to enforce a downward trend.

### 4.2 Bayesian Inference Procedure

More generally, we can express our goals-setting function as

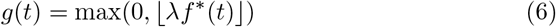

where *λ* is the user’s *learning rate* – the rate at which he or she can successfully reduce nicotine intake. The learning rate can be somewhat interpreted as the strength of the user’s addiction. If *λ* is too low, the goals are unachievable and the user is prone to relapse. On the other hand, if *λ* is too high, progress is minimal the cessation process is unnecessarily prolonged. In some sense, our learning rate here is analogous to the learning rate in gradient descent (usually denoted *α*), which determines the step size in each iteration. Each user has a unique learning rate that might even change throughout the cessation attempt, and thus we are tasked with learning the optimal *λ* over time.

We claim that *λ* can be learned using Bayesian inference. On day *t* = 0 we begin with some prior distribution, say *λ* ∼ 𝒩 (0.9, 0.2). Then for all days *t >* 0, we update our distribution with an iteration of Bayes’ theorem using the new evidence

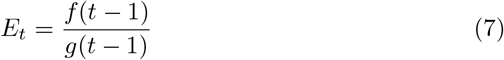

and we sample our new *λ* from the posterior distribution.

## 5 Conclusion

In this paper, we showed that Gaussian processes are an effective method for individualized modeling of nicotine addiction behaviors. In particular, GPs perform well when tasked with finding weekly patterns in a user’s smoking habits, and the outputted predictions for future usage are significantly more accurate than those of a naive forecasting model. We also proposed methods for leveraging the GP forecast outputs for goal-setting as part of a guided gradual cessation program. In future work, we intend to conduct a user study that validates the effectiveness of GPs in computer-guided smoking cessation.

## Data Availability

All data produced are available online at https://repositori.uji.es/xmlui/handle/10234/180682.

## Notes

### Competing Interest Statement

The authors have declared no competing interest.

### Funding Statement

This study did not receive any funding.

### Author Declarations

The study used only openly available human data that were originally located at https://repositori.uji.es/xmlui/handle/10234/180682.

